# Leveraging Expert Knowledge and Causal Structure Learning to Build Parsimonious Models of Acute Brain Dysfunction in the Pediatric Intensive Care Unit (PICU)

**DOI:** 10.64898/2026.02.17.26345661

**Authors:** Eddie Pérez Claudio, Christopher M. Horvat, W. Michael Taylor, Alicia K. Au, Robert S. B. Clark, Ruoting Li, Mehdi Nourelahi, Gregory F. Cooper, Harry Hochheiser

## Abstract

Machine learning adoption in clinical decision support systems remains limited by concerns about transparency and robustness. Causal structure learning (CSL) combined with expert knowledge may address these concerns by identifying potentially causal predictors, enabling more interpretable and clinically aligned models. In this study, we show that by integrating clinician expertise with CSL algorithms we can identify plausible causal drivers of acquired acute brain dysfunction (ABD) in the pediatric intensive care unit (PICU), which enables the development of parsimonious predictive models without substantial loss in performance. To do so, we analyzed 18,568 PICU encounters from the University of Pittsburgh Medical Center Children’s Hospital (2010–2022) and elicited knowledge from experienced clinicians. Encounters with acquired ABD were defined using the validated ABD computable phenotype. Expert knowledge was elicited from four clinicians through iterative interviews to construct a consensus directed acyclic graph (DAG). Clinician consensus achieved acceptable inter-rater reliability (Fleiss’ Kappa = 0.62) after two rounds of interviews and identified 16 biomarkers as potential causes of acquired ABD. Two CSL algorithms, GOLEM and PC-MB, were applied to enrich the clinician’s consensus DAG. The PC-MB algorithm showed 78% concordance with expert consensus, while GOLEM showed 46%. Together, the CSL algorithms identified seven biomarkers as potential causes that were not included in the clinician’s DAG: blood urea nitrogen, creatinine, dobutamine, glucose, potassium, PTT, SpO2. Using multiple variations of the enriched DAGs, XGBoost models were trained using biomarkers identified as potential causes of acquired ABD; these were evaluated primarily by area under the precision-recall curve (AUPRC). Models trained on the intersection of clinician consensus and PC-MB DAGs achieved an AUPRC of 0.79 (95% CI: 0.75–0.82) using only 14 biomarkers, compared to 0.81 (95% CI: 0.78–0.84) for the control model using all 45 biomarkers. When restricted to vitals and laboratory results alone, the best-performing model achieved an AUPRC of 0.77. Combining clinical expertise with causal structure learning enables the identification of causal hypotheses consistent with the clinical understanding of the participating clinicians and the development of parsimonious predictive models for acquired ABD in the PICU.

## Introduction

The widespread adoption of Machine Learning (ML) in clinical decision support systems (CDS) is hampered by concerns regarding trustworthiness [1–3]. Lack of insight into the reasoning behind the predictions ML models make [4–6] and their robustness to changes in clinical workflows [7,8] are central to these concerns. One possible approach for addressing these concerns involves the use of causally-informed predictive models that draw explicit links indicating how variables interact to produce the outcome [9–11]. When built using correct causal relationships, this class of models is more robust to out-of-distribution samples, meaning they have better performance on samples different from any data used for training [3,8]. When a ground-truth for causal knowledge is unavailable, as is often the case for clinical outcomes, causal structure learning (CSL) [12,13] methods may be used to approximate a plausible causal hypothesis, usually expressed in the form of a directed acyclic graph (DAG) [14,15]. These DAGs can then be used to guide the training of causally-informed predictive models. In this study, we build on our previous efforts in developing predictive models for acquired acute brain dysfunction (ABD) [16,17] to show that even with highly confounded EHR data, instantiating CSL methods with expert clinical knowledge can allow us to train causally-informed predictive models that are more transparent, robust, and consistent with clinician expertise when compared to more traditional correlation-based models.

Almost 40% of children admitted to the pediatric intensive care unit (PICU) acquire secondary brain injuries or related morbidities [18–20], encompassing a wide range of diagnoses, including altered consciousness, delirium, seizures, and encephalopathy [21,22], each with different causes and treatments. Although advances in critical care have reduced mortality associated with such injuries, acquired ABD has remained a major concern when trying to reduce the incidence of chronic neurological complications resulting from a stay in the PICU [20,23]. To reduce the harms from acquired ABD, clinicians need to identify at-risk patients as early as possible. Computable phenotypes have paved the way for our best efforts at improving the timeliness of risk identification [16,17,24,25]. Currently, the most accurate method to find patients with acquired ABD using electronic health record (EHR) data is the *Acute Brain Dysfunction* (ABD) computable phenotype [26–29]. The computable ABD phenotype has been validated through multiple chart reviews and achieves superior sensitivity and specificity compared to more established phenotypes based on the pupillary reaction of the patient or the Glasgow Coma Scale Scores (GCSS) [26]. Causal structure learning (CSL) techniques could be applied to longitudinal EHR data labeled with the ABD phenotype to identify vitals, medications, laboratory results, or procedures that may be causal drivers of acquired ABD in the PICU.

CSL, also known as causal search [29,30] or causal discovery [31,32], enables the inference of causal relationships from observational data under precisely specified assumptions [33,34]. While these methods generally learn only a subset of the causal relationships among a set of measured variables, the subset may include valuable and even novel causal relationships that are valid. Constraint-based methods like the Peter-Clark (PC) algorithm [12] estimate causal structures by performing a two-step process. The first step is to use conditional independence tests to identify an undirected graph. The second step applies rules to orient the edges in the graph and output a causal model in the form of a directed acyclic graph (DAG) [33,34]. Recent CSL algorithms like NOTEARS [35] or GOLEM [36] address this scaling issue by re-interpreting structure learning as a continuous linear optimization problem. While promising, the validity of the DAGs estimated by CSL methods can be limited by unmeasured confounders, selection bias, errors in the data. Some of these limitations might be mitigated through the inclusion of expert knowledge to augment purely data-driven structure learning [11,31,32].

When building a predictive model, the complexity of the CSL step can be reduced by focusing on identifying only the Markov Blanket (MB) for a target node [37–39]. In the current study, our target node is a*cquired ABD* in the PICU. MBs are the set of variables or nodes in a DAG that when conditioned upon make a target node independent of every other node; this includes the immediate ancestors of a node and its children [37]. Thus, with this MB, we can build causal ML models using XGBoost (XGB) [40] or other machine learning algorithms. Such causally-informed models have the potential to be achieve similar accuracy to fully specified models while using a smaller subset of the data [41].

We hypothesize that the ABD computable phenotype could be applied to a combination of longitudinal Electronic Health Record (EHR) data and expert knowledge to identify causal relationships across a patient’s vitals, labs, and medication regimen that lead to acquired ABD in the PICU. Using only the biomarkers inferred to be in the MB for ABD, we seek to build causally-informed predictive models that are more parsimonious than models containing all available biomarkers, but without drastic drops in model performance. We hypothesize that causally-informed models will be more understandable to clinicians than models that represent statistical associations that may not be causal. We also hypothesize that such causally-informed models will predict the acquired ABD outcome as well as models based on statistical association.

To test these hypotheses, we initialized a model with expert-specified knowledge of the causes of acquired ABD. To augment this model, we applied a CSL algorithm to EHR dataset containing 45 clinical biomarkers previously identified as having some degree of association with the acquisition of ABD in the PICU [17]. We assessed the validity of the estimated graphical causal models by showing consistency with the expert knowledge of our clinician collaborators (CH, RC, AU, MT). After removing potentially confounding biomarkers, the causally-informed and augmented predictive model had many fewer than 45 biomarkers, and yet it predicted acquired ABD almost as well as a model containing all 45 biomarkers. Our findings open the door to planned prospective studies for causally-informed risk models of ABD in the PICU and for applications of this approach in other clinical domain areas.

## Methods

To carry out our study, we applied the ABD computable phenotype to longitudinal EHR data from UPMC Children’s hospital. Specific windows of data before the ABD event were featurized and used to power the CSL and machine learning model training. Before running the CSL algorithms, we elicited knowledge on ABD from a group of PICU clinicians. Their insights were encoded as a DAG which we used as background knowledge for the CSL algorithms. Finally, we assessed which of the DAGs estimated by the CSL algorithms and their combinations with the clinician’s DAG, resulted in the best performing predictive model.

### Computable Phenotype for Acute Brain Dysfunction

We used the computable phenotype for ABD developed by Alcamo et al. (2020) [27] (Figure 1) to compute our target outcome. Patients who had any order for neurological imaging (computed tomography or magnetic resonance imaging), any behavioral health consultation, or any new anti-delirium medication were labeled as acquiring ABD.

**Figure 1.**
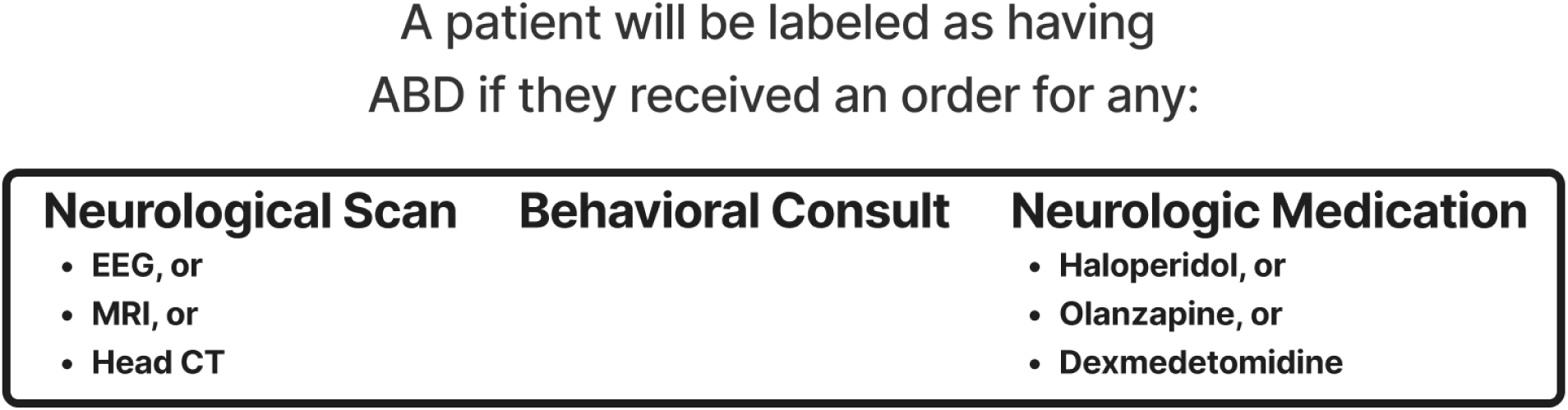
Computable Phenotype for ABD in the PICU

### Dataset

Our dataset contains 18,568 patient encounters from the Pediatric Intensive Care Unit (PICU) of the University of Pittsburgh Medical Center Children’s Hospital, occurring between January 1, 2010 and December 31, 2022. Each encounter has 45 biomarkers and associated demographic information. These biomarkers include vital signs, procedures, lab reports, and administered drugs. Like in our previous study [16], we scanned the longitudinal EHR data to find timepoints which coincided with the ABD phenotype. Once we identified an ABD event, we would censor, meaning we would exclude the twelve (12) hours right before the event. Our goal by censoring this data was to create a buffer. The ABD computable phenotype relies on orders for tests and drugs to estimate when event happened, these orders can sometimes show up in the EHR after a delay. Therefore, by censoring some of the data right before the event, we can reduce the chances we are including data that occurred after a clinician began suspecting the patient of ABD. To train our models we then used the 48 hours of data preceding the censored data. In our previous study [16], we tested different data windows; 48 hours provided us the best balance between model performance and sample capture, since many patients didn’t have much more than 48 hours of data after the censor window (Figure 2)

**Figure 2.**
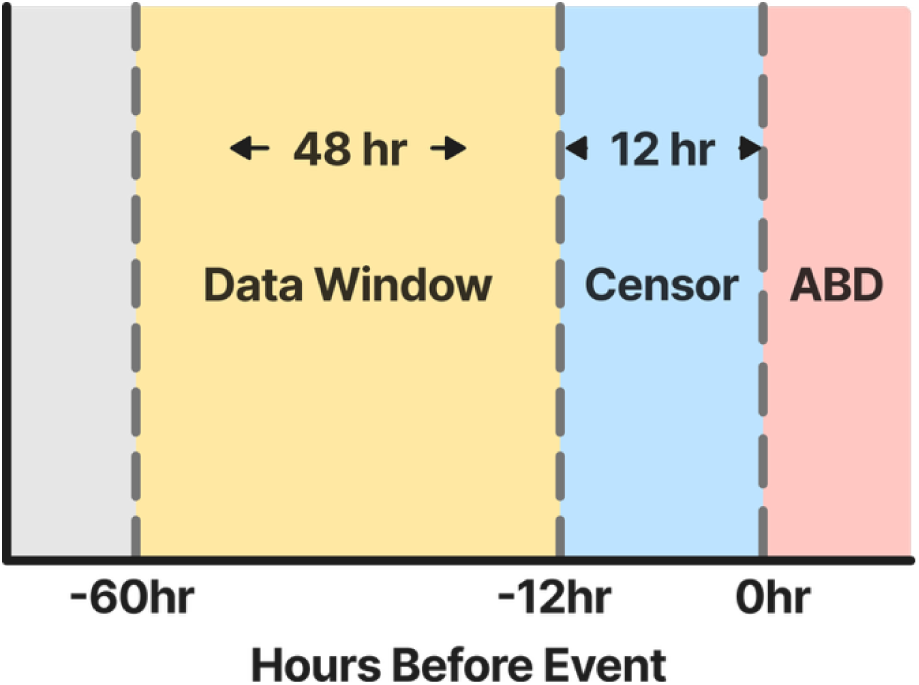
Data selection for featurization. Grey represents the time points before the window of interest. Yellow represents the 48-hour data window to be featurized. Data from this time window will be used in the models. Blue represents the censored time-points right before the ABD event. Red represents the data after the ABD event; this data is not included.

Before performing causal structure learning (CSL), we imputed missing values and featurized the biomarkers in the longitudinal EHR. The medications in our data set were encoded as binary variables and only the administration of a medication was recorded. Therefore, we imputed missing values with 0 by assuming that missing values indicate that the medication was not administered during that time-point. For Pupillary Reaction and other categorical variables, we performed a carry-forward imputation. Continuous variables were imputed using SAITS [42], a state-of-the-art model designed to impute data in partially observed multivariate timeseries. After imputing the missing values, we featurized the biomarkers in the longitudinal EHR data with Catch-22 [43],which extracts characteristics such as the shape of the distributions, the timing of extreme events, autocorrelation, and local trends for each biomarker. The featurization of our longitudinal EHR data led to a total of 990 features describing our 45 biomarkers over a 48-hour period.

We split the data into a training set and a testing set by year. Patient encounters from 2010 through 2019 were used for the training set (13,054 samples) and encounters from 2020 through 2022 were used as the testing set (3,895 samples). 13.54% of the train set consists of the positive class, our target acquired ABD outcome. 11.07% percent of the test set consists of the positive class.

We generated two (2) modified versions of the training set to ensure that we were taking the maximum advantage of the CSL algorithms we used while ensuring that they could run successfully in our computing environment (*Figure 3*). For Gradient-based optimization of DAG-penalized likelihood for linear dag models (GOLEM), a gradient based CSL algorithm which enables us to use large datasets, we removed collinear features (Pearson correlation *≥* 0.95) and low-variance features (variance *≤* 0.001) from the search set since such features would not add any significant amount of new information and could violate non-collinearity assumptions of the CSL algorithms. For the constraint-based algorithm, Peter-Clark Markov Blanket, which can grow in complexity with the density of the graph, we limited the search set to the top 3 features for each biomarker that were most correlated with the ABD target; since minimizing the number of features that come from the same biomarkers will help reduce the number of conditional independence tests to be carried out per biomarker.

**Figure 3.**
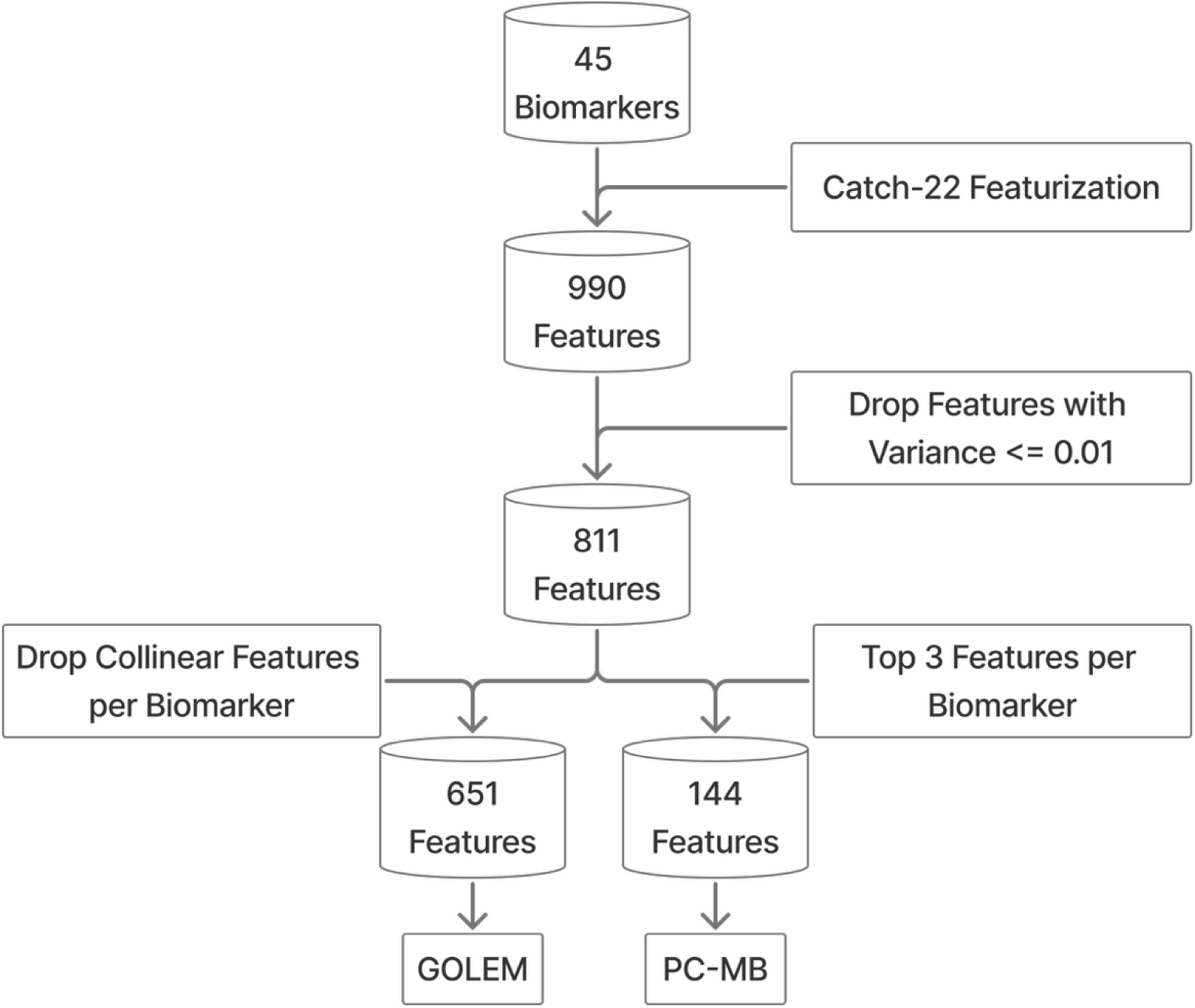
Data Processing Workflow.

### Expert Knowledge Elicitation

Given that we lack a ground truth for the causes of acquired PICU ABD, we established a working causal hypothesis based on clinical expertise. Expert knowledge was obtained through online interviews with four (4) senior PICU clinicians who specialize in neurocritical care (CH, AA, RC, and MT). The clinicians were given an interactive diagram containing 45 biomarkers associated with acquired brain injuries [14–18,20,21] (*Figure 4*). The goal was for them to draw a Directed Acyclic Graph (DAG) containing edges only from biomarkers they considered to be parents or causes to components of the computable ABD phenotype. Clinicians were asked to, within the context of ABD, add an edge to the DAG if an abnormal value for that biomarker or a set of biomarkers would cause them to suspect a patient is at risk of developing a brain injury or if an abnormal value would cause them to order an EEG, Brain MRI, Head CT, Behavioral Consult, or new neurologic medication. The DAGs of individual participants were joined into a consensus graph by including just those edges that appeared in at least 3 of 4 DAGs. We iterated through this process twice. During the second iteration, we used the consensus DAG as a starting from which we asked the four clinicians to add or remove edges. The final consensus DAG includes just those edges that appear in at least 3 of 4 DAGs.

**Figure 4.**
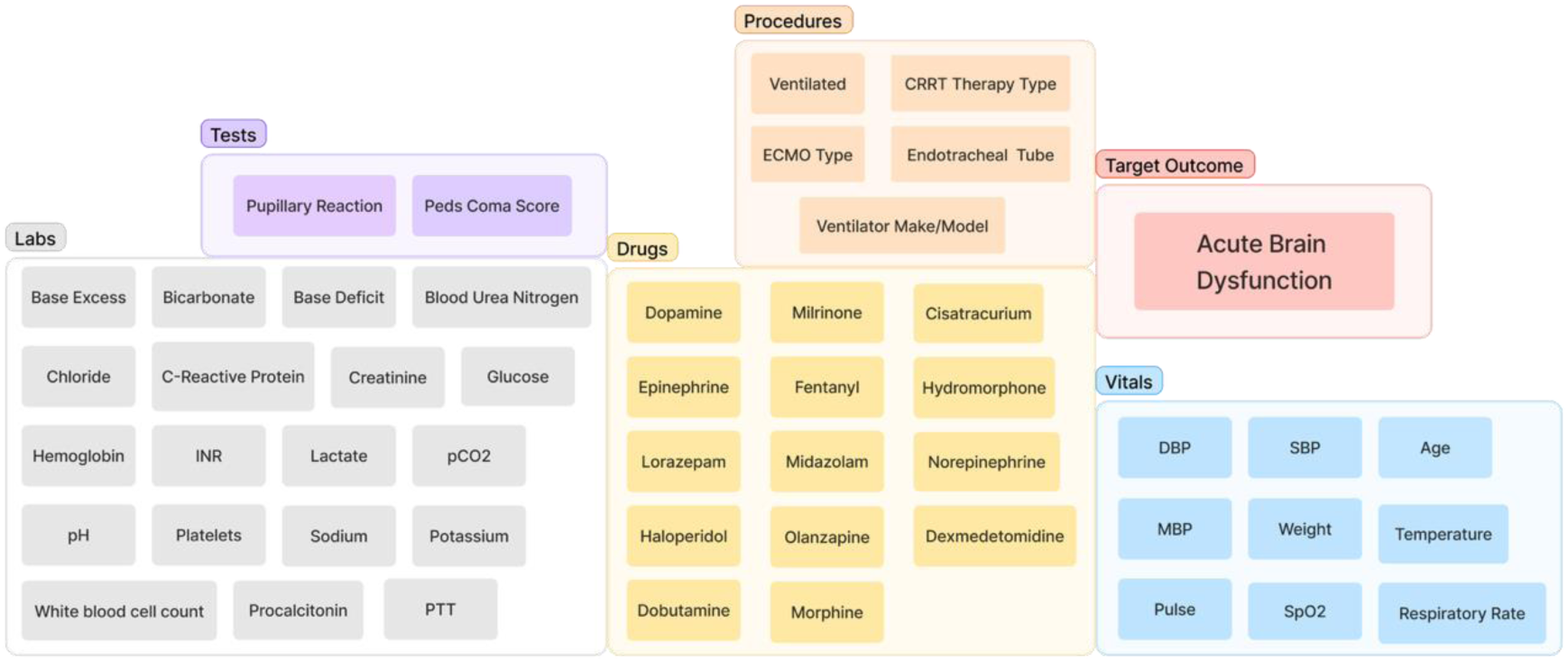
Biomarkers shown to participating clinicians with the target outcome.

To measure multi-rater agreement during each iteration we used Fleiss’ Kappa [44] as implemented in the Python library Statsmodels 0.14.2 [45]. By our second iteration we achieved a kappa score greater than 0.6, which is considered a measure of substantial agreement [46].

### Causal Structure Learning

#### Algorithm Description

CSL was performed using two algorithms. The first algorithm, Gradient-based optimization of DAG-penalized likelihood for linear dag models (GOLEM) [36], is based on novel neural network and gradient-based approaches to CSL. To estimate linear causal relationships, expert knowledge from clinicians was provided to the algorithm in the form of an adjacency matrix. We used the implementation of the GOLEM algorithm found in the gCASTLE repository [47]. The second algorithm was a variation of the classic Peter-Clark (PC) algorithm [12] that focuses on estimating the Markov Blanket (MB) of a particular target through conditional independence tests. The TETRAD 7.6.5 version [32] of the PC-MB algorithm was used.

#### Background Knowledge

Our best causal hypothesis for ABD in the PICU is based on the DAG built through expert consensus (Figure 5). We used this expert consenslus DAG as background knowledge to instantiate the CSL algorithms. For GOLEM, we transformed the expert consensus DAG into an adjacency matrix, where the presence or absence of an edge was encoded through binary indicators. One (1) implied the presence of an edge in the background knowledge and Zero (0) indicated the absence. This adjacency matrix skews the initial weights of the model so that it boosts the presence of the edges in the expert consensus DAG. For PC-MB we transformed the expert consensus DAG into a required edge list as described in the TETRAD 7.6.5 manual. This edge list was uploaded to the “Knowledge Box” in TETRAD and through visual inspection we verified that all edges matched the expert consensus. We did not add forbidden edges to the PC-MB background knowledge or add restrictions to the features the algorithms could use to learn the DAGs.

**Figure 5.**
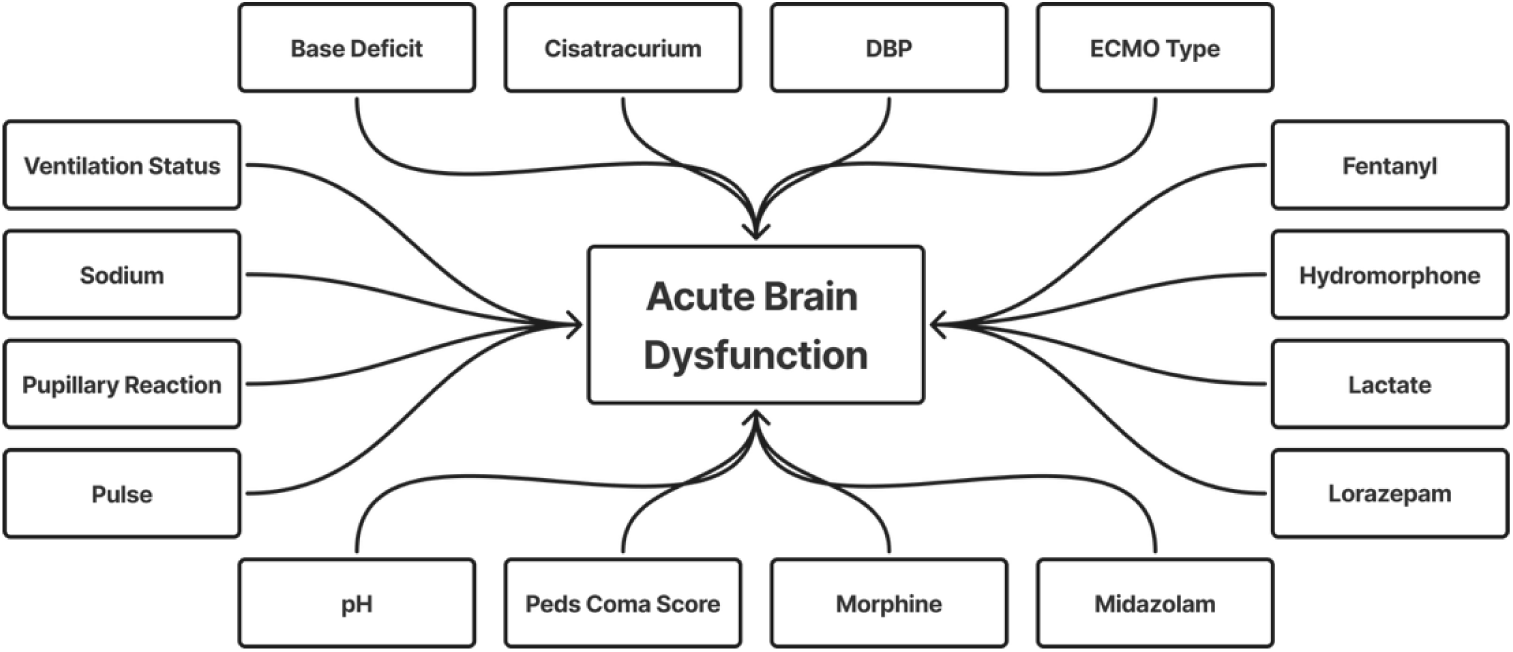
Simplified DAG from clinician consensus in the second iteration. In this version higher order interactions between biomarkers or components of the computable phenotype are not drawn. See Supplemental Figure 1 for the full DAG.

#### Enrichment of Clinician’s Consensus DAG

After the causal search, we used the expert consensus DAG as a base that we enriched in a data driven manner, with the information learned from the GOLEM [36] and PC-MB [12,32] algorithms (Figure 5). In particular, we generated modified versions of the clinicians’ consensus DAG which were either a union with the DAGs estimated by GOLEM, PC-MB, or an intersection of them where only the edges in common were kept.

### Predictive Models

#### Experiments

We performed three training experiments. In a previous study, Horvat et al 2025 [16], we built a predictive XGBoost model for ABD using the same dataset as in this study. However, we used a featurization approach which did not capture periodic or signal properties in the longitudinal EHR data. In the first experiment we tested whether a model using the same data, but a different featurization approach which can capture those signal properties can outperform our previous model. In this case, we used Catch22 [43] as the featurization approach. We compared the results of our previous paper against a “Control” model trained with the same biomarkers.

For our second experiment, we assessed whether a correlation-based or a causally-informed biomarker selection approach yields the best model for our dataset. To test this, we built a predictive model for each DAG identified in the CSL step, including the enriched variants of the clinician’s consensus DAG. The models were trained only the MB for each of the DAGs. For our dataset, the MB is comprised only of the parents for ABD (our target). This is because we only observe a 48-hour time window where we ensure the last timepoint is before the ABD target. By formatting the dataset in this way, we ensured the MB of ABD should be composed only of its parents as our target should have no child nodes. The “Correlation” model was trained using any biomarker whose features had a |Pearson correlation| ≥ 0.3 with the ABD target outcome.

Discussions with clinicians suggested that medications and other clinician actions are potential confounders, acting both as signals of and as responses to possible ABD. Thus, we performed a third experiment to assess the impact on model performance of removing these potential confounders from the training dataset, leaving only observational biomarkers like vitals and laboratory results.

#### Training

For each experiment, we trained a Linear Gaussian Bayesian Network (LGBN) [48] and an XGBoost [40] model as predictive models of the outcome. The structure of each LGBN followed the same structure as the DAG being parameterized. This way, LGBNs were used to explicitly encode linear interactions across the biomarkers predicting ABD. With XGBoost, we are able to model non-linear interactions between the biomarkers and ABD, but we cannot explicitly control which higher order interactions we will model across the biomarkers being used as predictors. All models were trained using only features estimated to be direct causes of ABD; therefore, resulting models function as structural equation models (SEM) which are commonly used to model causal relationships [38,39].

#### Testing

To evaluate model performance we used the following metrics: Accuracy, F1-Score, Precision, Recall, Accuracy and Area Under the Receiver Operating Characteristic Curver (AUROC), Area Under the Precision-Recall Curve (AUPRC) [49], and Expected Calibration Error (ECE) [50,51]. We calculated these metrics using the implementation of ECE in the Pycalib 0.1.0 [52] python library and the MLStatkit 0.1.91 [53] python library for all other metrics. The MLStatkit library enabled us to calculate the 95% Confidence Intervals (CI) for all metrics except ECE.

Due to the high class-imbalance in our dataset, traditional metrics like the Accuracy and AUROC could be overly optimistic [49]. Therefore, to assess model performance we focused on AUPRC which is more robust to class imbalance in the dataset and it provides us better insight at how well the model can distinguish between the binary objective (ABD = 1 vs ABD = 0) [49]. When multiple models had a similar AUPRC, we chose the most parsimonious model, defined as the model that used the fewest biomarkers, as the better performing model. We used AUROC as final tie breaker. We measured ECE to assess whether the class probabilities outputted by our models were properly calibrated [50,51].

#### Feature Importance

We evaluated the feature salience of the XGBoost models using the Tree-SHAP algorithm [54] from the Shapley Additive Explanations (SHAP) [55] python library. In the supplemental Figures 4-6, we show these at the biomarker level by calculating average absolute salience for each biomarker.

#### Code and Data Availability

Code to replicate this study can be found on GitHub: https://github.com/edpclau/casual_search_neuromorbidity. In addition, you will be able to find all supplementary material, hi-resolution files for all figures, and the step-by-step analysis performed to calculate multi-rater reliability.

We are unable to share the patient data used to train our models. However, the DAGs created by clinicians are available in the repository.

#### Institutional Review Board

The approval for this study was granted by the institutional review board of the University of Pittsburgh (STUDY20050220 and STUDY21090209).

## Results

### Expert Knowledge Elicitation

The first round of expert elicitations resulted in a Fleiss’ Kappa [44] of 0.195, reflecting poor alignment among the participating clinicians [46]. The second round of iteration resulted in a Fleiss’ Kappa of 0.62. This increase of 0.425 indicates a good level of reliability among clinicians. The consensus graph generated from the input of the clinicians contains 16 biomarkers. Out of these biomarkers, six (6) are laboratory results or tests, six (6) are medications, two (2) are indicators of a procedure, and two (2) are vitals (Figure 5). The biomarkers included in this clinical consensus DAG were in agreement with critical care literature [15,56–58].

### Causal Structure Learning

The GOLEM algorithm estimated 17 features to be direct parents of our computable target for ABD (Figure 6). These 17 features were derived using Catch22 from 11 unique biomarkers (**Error! Reference source not found.**). The PC-MB algorithm estimated 36 features to be direct causes of our computable target for ABD. Those 36 features are mapped to 17 unique biomarkers (Figure 7). For a list of all features, see Table S1.

**Figure 6.**
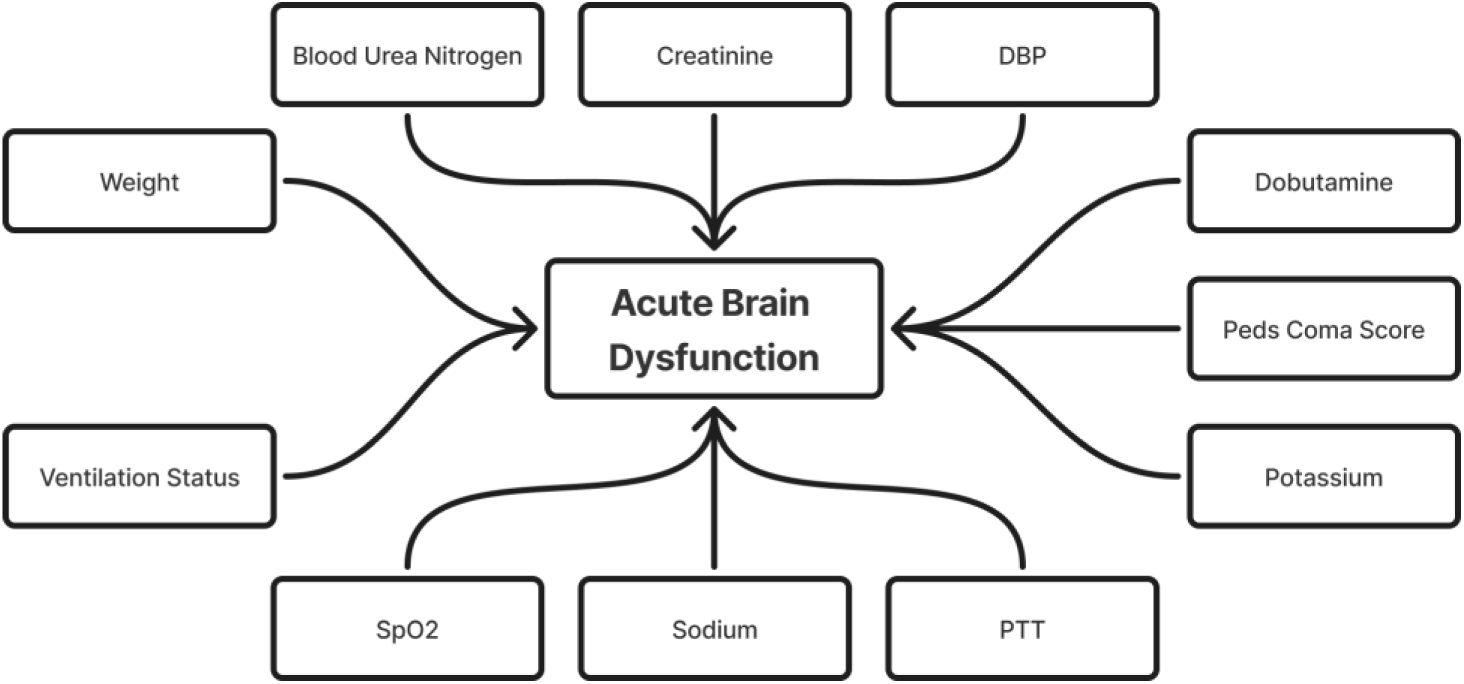
Simplified view of the proximal causes of ABD estimated by the GOLEM algorithms. Lines represent the directionality of likely causes. For more complex version of these DAGs see Supplementary Figure 2.

**Figure 7.**
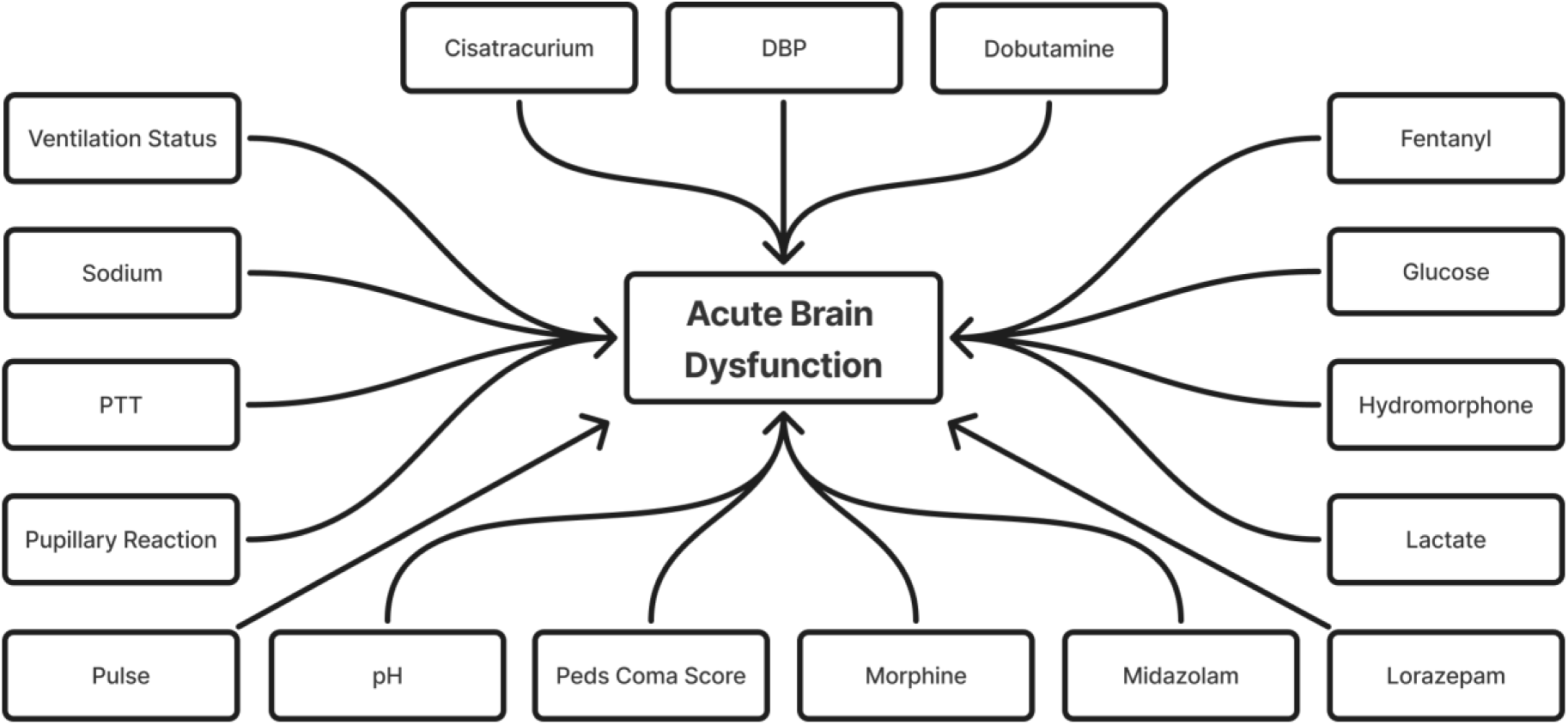
Simplified view of the proximal causes of ABD estimated by the PC-MB algorithm. Lines represent the directionality of likely causes. For more complex version of these DAGs see Supplementary Figure 3.

#### Face Validity

The face validity of the *GOLEM* and *PC-MB* DAGs (*Figure 6* and *Figure 7*) was evaluated by comparisons with the clinician consensus DAG (*Figure 5*). Of the 11 biomarkers included in the *GOLEM DAG*, five (46%) were also included in the clinician consensus DAG (*Table 1*).Table 1. Biomarkers in each DAG. X indicates the presence of a biomarker. Of the 18 biomarkers in the *PC-MB DAG*, 14 (78%) were also in the clinician consensus DAG. Of the 17 biomarkers in the clinician consensus DAG, 5 (29%) were also in the GOLEM DAG, and 15 (88%) were also in the PC-MB DAG. A participating clinician (CH) reviewed the biomarkers that were not present in the clinician consensus DAG and agreed that Blood Urea Nitrogen, Creatinine, Dobutamine, Potassium, PTT, Peripheral Oxygen Saturation (SPO2), and Glucose are clinically plausible biomarkers for acquired ABD. This is in line with observations in the critical care literature, where each of these biomarkers has been associated with ABD and neurological complications generally [57–63].

**Table 1.** Biomarkers in each DAG. X indicates the presence of a biomarker.

| **Biomarker** | **Clinician Consensus** | **PC-MB** | **GOLEM** |
| --- | --- | --- | --- |
| ABD (Outcome) | X | X | X |
| Base Deficit | X |  |  |
| Blood Urea Nitrogen |  |  | X |
| Cisatracurium | X | X |  |
| Creatinine |  |  | X |
| DBP | X | X | X |
| Dobutamine |  | X |  |
| Dopamine |  |  | X |
| ECMO Type | X |  |  |
| Fentanyl | X | X |  |
| Glucose |  | X |  |
| Hydromorphone | X | X |  |
| Lactate | X | X |  |
| Lorazepam | X | X |  |
| Midazolam | X | X |  |
| Morphine |  |  |  |
| Peds Coma Score | X | X | X |
| pH | X | X |  |
| Potassium |  |  | X |
| PTT | X |  | X |
| Pulse | X | X |  |
| Pupillary Reaction | X | X |  |
| Sodium | X | X | X |
| SpO2 |  |  | X |
| Ventilation Status | X | X | X |
| Weight |  |  | X |

Upon further review of these results, two of the participating clinicians (CH and AA) suggested removing Ventilation Status from all DAGs, since it is a highly confounded indicator of patient deterioration. They also suggested removing Extra Corporeal Membrane Oxygenation (ECMO) Type from the clinician consensus DAG since this is also a highly confounded indicator of patient deterioration.

#### Enrichment of Clinician’s Consensus DAG

We enriched the *clinician consensus* DAG through union and intersection with the estimated DAGs (*Table 2*). The *clinician consensus ∪ GOLEM* DAG has six (6) additional biomarkers (*Table 1*). The *clinician consensus ∪ PC-MB* DAG has four (4) additional biomarkers. The *clinician consensus ∩ GOLEM* DAG has four (4) biomarkers, if we exclude Ventilation Status. The *clinician consensus ∩ PC-MB* DAG is similar to the *clinician consensus* DAG: only Base Deficit is missing. The *PC-MB ∪ GOLEM*

**Table 2.** Enrichment Strategies

| DAG | Enrichment Strategy | Biomarkers | Change from Baseline |
| --- | --- | --- | --- |
| Clinician Consensus ∪ PC-MB | Union | 18 | +3 |
| Clinician Consensus ∪ GOLEM | Union | 22 | +7 |
| PC-MB ∪ GOLEM | Union | 23 | +6 |
| Clinician Consensus ∩ PC-MB | Intersection | 14 | -1 |
| Clinician Consensus ∩ GOLEM | Intersection | 4 | -11 |
| PC-MB ∩ GOLEM | Intersection | 5 | -13 |

DAG has 23 biomarkers, only four (4) biomarkers were in common: Diastolic Blood Pressure (DBP), Peds Coma Score, Partial Thromboplastin Time (PTT), and Sodium.

### Predictive Model Performance

#### Changing the featurization approach to Catch22 provided our predictive models a significant boost in performance

In Horvat et al (2025), we trained an XGBoost model using the same dataset and target to achieve and AUPRC of 0.62 (95% 0.58, 0.63) (*Table 3*). By changing the featurization approach to Catch22, the control model in the current study achieves an AUPRC of 0.81 (95% 0.78, 0.84), which is a 19% improvement.

**Table 3.**
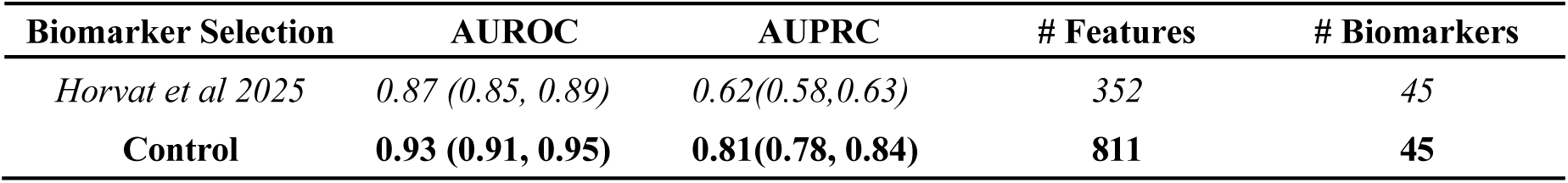
Results of changing the featurization approach to Catch22. The line in bold represents the model with the better performance across AUROC and AUPRC.

#### Expert knowledge with CSL outperforms correlation-based biomarker selection methods while using fewer biomarkers

Out of all the experimental models, the XGBoost model trained with the biomarkers on the *clinician consensus ∩ PC-MB* DAG (bolded in *Table 4*) had the best performance with an AUPRC of 0.79 (95%CI 0.75, 0.82). Which is only a 2% reduction when compared to the Control model. However, this performance was achieved with 14 biomarkers, 31 (69%) fewer than the Control model. The XGBoost model based on the *clinician consensus ∩ PC-MB* DAG also outperformed the Correlation model by 4% with 23 (62%) fewer biomarkers, showing the advantage of using the MB identified through expert knowledge and CSL for biomarker selection over correlational associations.

**Table 4.** Results of Biomarker Selection Experiments. The line in bold represents the experimental model that best balances AUPRC performance and biomarker parsimony.

| Biomarker Selection | AUROC | AUPRC | Precision | Recall | F1 Score | Accuracy | ECE | # Features | # Biomarkers |
| --- | --- | --- | --- | --- | --- | --- | --- | --- | --- |
| Control | 0.93 (0.91, 0.95) | 0.81 (0.78, 0.84) | 0.94 (0.92, 0.95) | 0.80 (0.78, 0.83) | 0.85 (0.83, 0.87) | 0.95 (0.94, 0.96) | 0.089 | 811 | 45 |
| Correlation Feature Selection | 0.90 (0.88, 0.91)* | 0.75 (0.71, 0.78) | 0.90 (0.88, 0.92) | 0.78 (0.76, 0.81) | 0.83 (0.81, 0.85) | 0.94 (0.94, 0.95) | 0.108 | 113 | 37 |
| Clinician Consensus | 0.91 (0.89, 0.93)* | 0.77 (0.74, 0.81) | 0.92 (0.90, 0.94) | 0.79 (0.77, 0.82) | 0.84 (0.82, 0.86) | 0.95 (0.94, 0.95) | 0.095 | 259 | 15 |
| GOLEM | 0.91 (0.89, 0.93)* | 0.77 (0.74, 0.80) | 0.91 (0.90, 0.93) | 0.79 (0.76, 0.81) | 0.83 (0.82, 0.86) | 0.94 (0.94, 0.95) | 0.095 | 199 | 11 |
| PC-MB | 0.91 (0.89, 0.93)* | 0.78 (0.75, 0.82) | 0.92 (0.91, 0.94) | 0.79 (0.77, 0.81) | 0.84 (0.82, 0.86) | 0.95 (0.94, 0.95) | 0.092 | 296 | 17 |
| Clinician Consensus ∪ GOLEM | 0.91 (0.89, 0.93)* | 0.78 (0.75, 0.82) | 0.92 (0.90, 0.94) | 0.79 (0.77, 0.82) | 0.84 (0.82, 0.86) | 0.95 (0.94, 0.95) | 0.092 | 389 | 22 |
| Clinician Consensus ∪ PC-MB | 0.91 (0.89, 0.93)* | 0.79 (0.75, 0.82) | 0.92 (0.90, 0.93) | 0.80 (0.78, 0.83) | 0.85 (0.83, 0.87) | 0.95 (0.94, 0.96) | 0.094 | 315 | 18 |
| GOLEM ∪ PC-MB | 0.91 (0.89, 0.93)* | 0.78 (0.75, 0.81) | 0.91 (0.89, 0.93) | 0.79 (0.77, 0.81) | 0.84 (0.82, 0.86) | 0.94 (0.94, 0.95) | 0.093 | 407 | 23 |
| Clinician Consensus ∩ GOLEM | 0.89 (0.87, 0.91)* | 0.74 (0.70, 0.77) | 0.89 (0.87, 0.91) | 0.78 (0.76, 0.80) | 0.82 (0.80, 0.84) | 0.94 (0.93, 0.95) | 0.106 | 69 | 4 |
| **Clinician Consensus ∩ PC-MB** | **0.91 (0.89, 0.93)\*** | **0.79 (0.75, 0.82)** | **0.93 (0.91, 0.94)** | **0.80 (0.77, 0.82)** | **0.85 (0.82, 0.86)** | **0.95 (0.94, 0.96)** | **0.095** | **240** | **14** |
| GOLEM ∩ PC-MB | 0.90 (0.89, 0.92)* | 0.76 (0.73, 0.80) | 0.90 (0.88, 0.92) | 0.79 (0.77, 0.81) | 0.83 (0.81, 0.85) | 0.94 (0.94, 0.95) | 0.102 | 88 | 5 |

\* : AUROC is significantly different from the Control.

#### Simplified models based solely on vitals and laboratory results had comparable performance with far fewer biomarkers

In our last experiment, we kept only biomarkers related to vitals and laboratory results in our datasets. Under this condition, the Control had a total of 28 biomarkers and an AUPRC of 0.78 (95%CI 0.75, 0.82), a 3% reduction from using the full dataset (*Table 4* and *Table 5*). Under these conditions, the XGBoost model based on the *clinician consensus ∪ GOLEM DAG had an AUPRC of 0.77 (95%CI 0.74, 0.81), which is a 1% decrease from the Control model (Table 5) while using 17 (61%) fewer biomarkers*.

**Table 5.** Model Performance with reduced feature set containing only vitals and laboratory results. The line in bold represents the experimental model that best balances AUPRC performance and biomarker parsimony.

| Biomarker Selection | AUROC | AUPRC | Precision | Recall | F1 Score | Accuracy | ECE | # Features | # Biomarkers |
| --- | --- | --- | --- | --- | --- | --- | --- | --- | --- |
| Control | 0.92 (0.90, 0.93) | 0.78 (0.75, 0.82) | 0.90 (0.88, 0.92) | 0.80 (0.78, 0.82) | 0.84 (0.82, 0.86) | 0.95 (0.94, 0.95) | 0.093 | 521 | 28 |
| Correlation Feature Selection | 0.89 (0.87, 0.91)* | 0.75 (0.71, 0.78) | 0.90 (0.88, 0.92) | 0.78 (0.76, 0.80) | 0.83 (0.80, 0.85) | 0.94 (0.93, 0.95) | 0.114 | 53 | 20 |
| Clinician Consensus | 0.89 (0.87, 0.91)* | 0.73 (0.69, 0.76) | 0.89 (0.87, 0.91) | 0.78 (0.75, 0.80) | 0.82 (0.80, 0.84) | 0.94 (0.93, 0.95) | 0.108 | 93 | 5 |
| GOLEM | 0.91 (0.89, 0.92)* | 0.76 (0.72, 0.79) | 0.90 (0.88, 0.92) | 0.79 (0.77, 0.81) | 0.83 (0.81, 0.85) | 0.94 (0.94, 0.95) | 0.102 | 149 | 8 |
| PC-MB | 0.91 (0.89, 0.92)* | 0.76 (0.73, 0.80) | 0.89 (0.87, 0.91) | 0.79 (0.77, 0.81) | 0.83 (0.81, 0.85) | 0.94 (0.93, 0.95) | 0.104 | 131 | 7 |
| **Clinician Consensus ∪ GOLEM** | **0.91 (0.90, 0.93)** | **0.77 (0.74, 0.81)** | **0.91 (0.89, 0.93)** | **0.79 (0.77, 0.81)** | **0.84 (0.82, 0.86)** | **0.95 (0.94, 0.95)** | **0.101** | **204** | **11** |
| Clinician Consensus ∪ PC-MB | 0.91 (0.89, 0.92) | 0.76 (0.73, 0.80) | 0.89 (0.87, 0.91) | 0.79 (0.77, 0.81) | 0.83 (0.81, 0.85) | 0.94 (0.93, 0.95) | 0.104 | 131 | 7 |
| GOLEM ∪ PC-MB | 0.91 (0.89, 0.93) | 0.77 (0.73, 0.80) | 0.91 (0.89, 0.93) | 0.78 (0.76, 0.81) | 0.83 (0.81, 0.85) | 0.94 (0.94, 0.95) | 0.098 | 223 | 12 |
| Clinician Consensus ∩ GOLEM | 0.86 (0.84, 0.88)* | 0.66 (0.62, 0.70) | 0.85 (0.82, 0.87) | 0.74 (0.72, 0.76) | 0.78 (0.76, 0.80) | 0.93 (0.92, 0.93) | 0.116 | 38 | 2 |
| Clinician Consensus ∩ PC-MB | 0.89 (0.87, 0.91)* | 0.73 (0.69, 0.76) | 0.89 (0.87, 0.91) | 0.78 (0.75, 0.80) | 0.82 (0.80, 0.84) | 0.94 (0.93, 0.95) | 0.108 | 93 | 5 |
| GOLEM ∩ PC-MB | 0.89 (0.87, 0.91)* | 0.71 (0.68, 0.75) | 0.86 (0.84, 0.89) | 0.77 (0.75, 0.80) | 0.81 (0.79, 0.83) | 0.93 (0.93, 0.94) | 0.113 | 57 | 3 |

\* : AUROC is significantly different from the Control.

#### XGBoost based predictive models outperformed LGBNs in all experiments

During model testing, XGBoost outperformed the LGBNs regardless of the DAG they were based on. To see the LGBN results, see Supplementary Tables 3, 5, and 7.

## Discussion

### Overview

Transparency and robustness are critical requirements for clinical predictive models [4–6]. Using causal DAGs to select Markov Blanket (MB) variables for a predictive model is a promising approach for addressing these concerns [37,41]. Such predictive models can be more interpretable and more robust in the face of outliers [3]. Our study adds to this body of evidence [5,6,38,39,64–66], showing that the combination of clinical expertise and causal structure learning (CSL) enables the estimation of plausible MBs in EHR data to build more parsimonious and causality-aware predictive models. We investigated novel expert-driven and data-driven causal DAGs for pediatric acute brain dysfunction (ABD) in the PICU. The predictive models we trained using the MB of these DAGs achieved performance comparable to using all available predictive variables while using fewer, but potentially causal, biomarkers. Our approach reduces the risk that confounded or spuriously predictive variables are being used for making predictions. This is especially true for the models that we trained on strictly observational biomarkers (vitals and laboratory results), as the AUPRC difference between the best model and the control was less than one percent (1%). These results support our hypothesis that learning causal structures, when combined with expert knowledge, can allow us to build more parsimonious and causally-aware models with minimal, if any, impact on predictive performance.

### Causal Structure Learning (CSL)

Concepts of diverse pathologies, clinical outcomes such as ABD are difficult to define narrowly [22]. Efforts to predict or identify ABD rely on multi-factorial measurements and clinical judgment [16,18,67]. However, expert models may not be rigorously informed or incomplete and thus can exclude important information (*Figure 5*).

CSL is increasingly being used to develop data-driven clinical predictive models with great success in terms of improving predictive model robustness [3,11] by reducing the influence of spuriously predictive variables and model complexity by allowing us to predict using only the MB [41,68]. These CSL approaches can estimate causal interactions across hundreds of features on a scale that is difficult for human experts. Since exhaustive search is computationally intractable, CSL algorithms use heuristic approximations. These approximations are non-deterministic, meaning that in highly confounded datasets, the starting point of the search may lead to different DAGs. Incorporating expert judgment as background knowledge for these algorithms mitigates some of their limitations by narrowing the search space and setting starting point [69,70]. In our study, we also found that this combined approach can encourage clinicians to reflect on their experiences and biases, thus refining their mental models of disease.

We based our causal search on a robust DAG generated through multiple rounds of clinician interviews and focus groups. This initial clinical consensus DAG was in agreement with critical care literature [16,18,56–58] (*Figure 5*). After using the clinical DAG as background knowledge for the GOLEM and PC-MB algorithms, we identified Creatinine, Dobutamine, Glucose, Potassium, PTT, SpO2, and Blood Urea Nitrogen as potentially overlooked causes of acquired ABD in the PICU. Upon further exploration, we found that these seven (7) biomarkers are consistent with observations in critical care literature [58–63]. This implies these seven (7) biomarkers obtained through data-driven CSL algorithms may be valid causal candidates of ABD that the participating clinicians overlooked. When presented with this new information, some participating clinicians (CH, AA) reflected and indicated that they should have included these biomarkers in their expert-based DAG.

### Predictive Modeling

Previous models for ABD in the PICU [16,71], used a featurization approach which does not capture circadian or periodic/cyclic changes in a patient’s biomarkers. This limits how expressive the models can be since important time series properties of longitudinal EHR data is being missed. Switching our featurization approach to Catch-22 [43], which extracts such important information, we were able to improve the performance of the AUPRC of the model by 19% (*Table 3*). We leveraged this more expressive featurization approach and the causal DAGs we estimated in our CSL step to include only the biomarkers estimated to be potential causes ABD in the PICU. This allowed us to build a set of predictive models that, unlike previous ones, are causally-aware and use a reduced set of biomarkers which improves the model’s generalizability and utility. With the need for 34 (76%) fewer biomarkers over previous models [16], this new casually-aware predictive model (bolded in *Table 5*) has the potential to be applied to more patients, with fewer resources, and in more institutions. Furthermore, we did this with a subset of vitals and laboratory results free of clearly confounded biomarkers such as medications and procedures. An added benefit of this approach is that the biomarkers incorporated in the model are all part of the clinician’s mental model of acquired ABD and thus might be more likely to be used and understood by clinicians.

### Limitations and Future Work

The GOLEM and PC-MB algorithms assume causal sufficiency, meaning they do not model for the possibility of unmeasured, hidden confounders [12,36]. Therefore, our findings as to whether certain biomarkers are, or not, direct causes of acquired ABD should be taken as preliminary, as there is a possibility of unmeasured variables confounding those relationships [72]. As we gain access to more data and computing resources, we plan to expand the list of biomarkers we include in our causal search and use compute-intensive algorithms like Best Order Score Search - Fast Causal Inference (BOSS-FCI) [32,73,74], which can estimate the presence of potential hidden confounders.

A second limitation of our study is our use of a featurized dataset observing only a 48-hour window of data. This restricted view creates the possibility that our model would miss any direct cause of acquired ABD occurring before the 48-hour window. Therefore, the biomarkers we found to be direct parents of ABD are likely a subset of all possible causes. To minimize the loss of this information, we will explore the application of causal structure learning algorithms that handle timeseries data natively [75,76]. Searching over timeseries data will afford us the possibility of using the full dataset to find critical time points towards the development of acquired ABD.

As we work to overcome the limitations of our study, the next steps include incorporating clinical notes to refine our ABD outcome, developing a clinical decision support interface for our predictive models, and running the predictive models we have developed so far in a prospective study.

## Conclusion

Our work shows combining expert knowledge and causal structure learning can enable the estimation of causal DAGs for ABD in the PICU that have clinical face validity and concur with critical care literature. Furthermore, we present a novel list of potential causes of ABD in the PICU which includes: *Blood Urea Nitrogen, Creatinine, DBP, Dobutamine, Potassium, PTT, Sodium, and SpO2* as good candidates. However, further work will be needed to validate these biomarkers as causally influencing the occurrence of ABD and to model how they interact to produce ABD. The predictive models trained using these biomarkers show promise as potential tools to power causally-aware clinical decision support tools to manage the risk of acquired ABD in the PICU. The approach we used here can be used to develop causally-informed models for other healthcare domains.

## Data Availability

With the exception of patient data, all data produced in the present study are available upon reasonable request to the authors. The code to reproduce our study can be found in our github.

https://github.com/edpclau/CSL_ABD

## Acknowledgments and Funding

We acknowledge funding from NINDS R01NS118716, NIH 5T32HD040686-23, NLM/NIH 5T15LM007059-38, and NIH R01HL164835.

This research was supported in part by the University of Pittsburgh Center for Research Computing through the resources provided. Specifically, this work used the H2P cluster, which is supported by NSF award number OAC-2117681.

