## Supplemental Tables and Figures for "Leveraging Expert Knowledge and Causal Structure Learning to Build Parsimonious Models of Acute Brain Dysfunction in the Pediatric Intensive Care Unit (PICU)"

Supplementary Table 1. Features present in the GOLEM and PC-MB DAGs. See the official documentation 71 for an in-depth description of Catch22 features.

**Catch22	Counts**

**Category	Feature	GOLEM	PC-MB**

Distribution Shape	mode

10	5	-

Distribution Shape	mode 5	1	- Linear Autocorrelation	acf

timescale	-	4

Linear Autocorrelation	centroid freq	-	7

Linear Autocorrelation	low

freq power	2	1

Linear Autocorrelation Structure	periodicity	1	2

Symbolic	stetch

decreasing	-	10

Symbolic	transition

variance	-	4

Incremental Differences	whiten

timescale	-	2

Incremental Differences	high

fluctuation	-	1

Other	embedding

dist	3	-

Other	mean	3	-

Other	std	1	-

Supplementary Table 2. Results of Delong’s test 72 for XGBoost models in the Biomarker Selection Experiment.

| **Model 1** | **Model 2** | **Z-score** | **P-value** |
| --- | --- | --- | --- |
| Control | Correlation | -5.0469 | 4.491e-07 |
| Control | Clinician Consensus | -3.8218 | 1.325e-04 |
| Control | GOLEM | -3.1809 | 1.468e-03 |
| Control | PC-MB | -3.7981 | 1.458e-04 |
| Control | Clinician Consensus *∪* GOLEM | -3.5661 | 3.623e-04 |
| Control | Clinician Consensus *∪* PC-MB | -3.1917 | 1.415e-03 |
| Control | GOLEM *∪* PC-MB | -3.4346 | 5.934e-04 |
| Control | Clinician Consensus *∩* GOLEM | -5.5644 | 2.630e-08 |
| Control | Clinician Consensus *∩* PC-MB | -3.5869 | 3.346e-04 |
| Control | GOLEM *∩* PC-MB | -3.7163 | 2.022e-04 |

Supplementary Table 3. Full Results of Biomarker Selection Experiments

| **Model** | **DAG** | **AUPRC** | **AUROC** | **Precision** | **Recall** | **F1 Score** | **Accuracy** | **ECE** | **# Features** | **# Biomarkers** |
| --- | --- | --- | --- | --- | --- | --- | --- | --- | --- | --- |
| LGBN | Control | 0.76 (0.72, 0.80) | 0.90 (0.88, 0.92) | 0.91 (0.89, 0.93) | 0.78 (0.76, 0.81) | 0.83 (0.81, 0.85) | 0.94 (0.94, 0.95) | 0.139 | 811 | 45 |
| XGB | Control | 0.81 (0.78, 0.84) | 0.93 (0.91, 0.95) | 0.94 (0.92, 0.95) | 0.80 (0.78, 0.83) | 0.85 (0.83, 0.87) | 0.95 (0.94, 0.96) | 0.089 | 811 | 45 |
| LGBN | Correlation | 0.73 (0.69, 0.76) | 0.87 (0.84, 0.89) | 0.95 (0.93, 0.96) | 0.74 (0.72, 0.77) | 0.81 (0.79, 0.83) | 0.94 (0.93, 0.95) | 0.133 | 113 | 37 |
| XGB | Correlation | 0.75 (0.71, 0.78) | 0.90 (0.88, 0.91) | 0.90 (0.88, 0.92) | 0.78 (0.76, 0.81) | 0.83 (0.81, 0.85) | 0.94 (0.94, 0.95) | 0.108 | 113 | 37 |
| LGBN | Clinician Consensus *∪* PC-MB | 0.76 (0.72, 0.79) | 0.91 (0.89, 0.93) | 0.93 (0.92, 0.95) | 0.77 (0.74, 0.79) | 0.83 (0.80, 0.85) | 0.94 (0.94, 0.95) | 0.136 | 315 | 18 |
| XGB | Clinician Consensus *∪* PC-MB | 0.79 (0.75, 0.82) | 0.91 (0.89, 0.93) | 0.92 (0.90, 0.93) | 0.80 (0.78, 0.83) | 0.85 (0.83, 0.87) | 0.95 (0.94, 0.96) | 0.094 | 315 | 18 |
| LGBN | Clinician Consensus *∪* GOLEM | 0.74 (0.70, 0.78) | 0.90 (0.88, 0.92) | 0.93 (0.92, 0.95) | 0.77 (0.75, 0.79) | 0.83 (0.81, 0.85) | 0.94 (0.94, 0.95) | 0.14 | 389 | 22 |
| XGB | Clinician Consensus *∪* GOLEM | 0.78 (0.75, 0.82) | 0.91 (0.89, 0.93) | 0.92 (0.90, 0.94) | 0.79 (0.77, 0.82) | 0.84 (0.82, 0.86) | 0.95 (0.94, 0.95) | 0.092 | 389 | 22 |
| LGBN | GOLEM *∪* PC-MB | 0.75 (0.71, 0.79) | 0.91 (0.89, 0.92) | 0.93 (0.92, 0.95) | 0.77 (0.75, 0.80) | 0.83 (0.81, 0.85) | 0.94 (0.94, 0.95) | 0.141 | 407 | 23 |
| XGB | GOLEM *∪* PC-MB | 0.78 (0.75, 0.81) | 0.91 (0.89, 0.93) | 0.91 (0.89, 0.93) | 0.79 (0.77, 0.81) | 0.84 (0.82, 0.86) | 0.94 (0.94, 0.95) | 0.093 | 407 | 23 |
| LGBN | Clinician Consensus | 0.75 (0.72, 0.79) | 0.90 (0.88, 0.92) | 0.94 (0.93, 0.96) | 0.76 (0.74, 0.78) | 0.82 (0.80, 0.84) | 0.94 (0.94, 0.95) | 0.132 | 259 | 15 |
| XGB | Clinician Consensus | 0.77 (0.74, 0.81) | 0.91 (0.89, 0.93) | 0.92 (0.90, 0.94) | 0.79 (0.77, 0.82) | 0.84 (0.82, 0.86) | 0.95 (0.94, 0.95) | 0.095 | 259 | 15 |
| LGBN | Clinician Consensus *∩* PC-MB | 0.75 (0.72, 0.79) | 0.90 (0.88, 0.92) | 0.95 (0.93, 0.96) | 0.76 (0.74, 0.78) | 0.82 (0.80, 0.84) | 0.94 (0.94, 0.95) | 0.132 | 240 | 14 |
| XGB | Clinician Consensus *∩* PC-MB | 0.79 (0.75, 0.82) | 0.91 (0.89, 0.93) | 0.93 (0.91, 0.94) | 0.80 (0.77, 0.82) | 0.85 (0.82, 0.86) | 0.95 (0.94, 0.96) | 0.095 | 240 | 14 |
| LGBN | PC-MB | 0.76 (0.72, 0.79) | 0.91 (0.89, 0.92) | 0.94 (0.92, 0.95) | 0.77 (0.74, 0.79) | 0.83 (0.80, 0.85) | 0.94 (0.94, 0.95) | 0.136 | 296 | 17 |
| XGB | PC-MB | 0.78 (0.75, 0.82) | 0.91 (0.89, 0.93) | 0.92 (0.91, 0.94) | 0.79 (0.77, 0.81) | 0.84 (0.82, 0.86) | 0.95 (0.94, 0.95) | 0.092 | 296 | 17 |
| LGBN | GOLEM | 0.73 (0.70, 0.77) | 0.90 (0.88, 0.91) | 0.92 (0.90, 0.94) | 0.75 (0.73, 0.78) | 0.81 (0.79, 0.83) | 0.94 (0.93, 0.95) | 0.138 | 199 | 11 |
| XGB | GOLEM | 0.77 (0.74, 0.80) | 0.91 (0.89, 0.93) | 0.91 (0.90, 0.93) | 0.79 (0.76, 0.81) | 0.83 (0.82, 0.86) | 0.94 (0.94, 0.95) | 0.095 | 199 | 11 |
| LGBN | GOLEM *∩* PC-MB | 0.71 (0.67, 0.76) | 0.89 (0.87, 0.91) | 0.93 (0.91, 0.94) | 0.74 (0.71, 0.76) | 0.80 (0.77, 0.82) | 0.94 (0.93, 0.95) | 0.131 | 88 | 5 |
| XGB | GOLEM *∩* PC-MB | 0.76 (0.73, 0.80) | 0.90 (0.89, 0.92) | 0.90 (0.88, 0.92) | 0.79 (0.77, 0.81) | 0.83 (0.81, 0.85) | 0.94 (0.94, 0.95) | 0.102 | 88 | 5 |
| LGBN | Clinician Consensus *∩* GOLEM | 0.72 (0.67, 0.75) | 0.88 (0.86, 0.90) | 0.94 (0.92, 0.95) | 0.73 (0.70, 0.75) | 0.79 (0.77, 0.81) | 0.94 (0.93, 0.94) | 0.13 | 69 | 4 |
| XGB | Clinician Consensus *∩* GOLEM | 0.74 (0.70, 0.77) | 0.89 (0.87, 0.91) | 0.89 (0.87, 0.91) | 0.78 (0.76, 0.80) | 0.82 (0.80, 0.84) | 0.94 (0.93, 0.95) | 0.106 | 69 | 4 |

*Supplementary Table 4. Results of Delong’s test 72 for XGBoost models in the No Medications Experiment.*

| **Model 1** | **Model 2** | **Z-score** | **P-value** |
| --- | --- | --- | --- |
| Control | Correlation | -4.7039 | 2.553e-06 |
| Control | Clinician Consensus *∪* PC-MB | -2.8541 | 4.317e-03 |
| Control | Clinician Consensus *∪* GOLEM | -2.722 | 6.488e-03 |
| Control | GOLEM *∪* PC-MB | -2.6067 | 9.141e-03 |
| Control | Clinician Consensus | -3.912 | 9.154e-05 |
| Control | Clinician Consensus *∩* PC-MB | -3.0183 | 2.542e-03 |
| Control | PC-MB | -2.2769 | 2.279e-02 |
| Control | GOLEM | -1.0311 | 3.025e-01 |
| Control | GOLEM *∩* PC-MB | -2.8589 | 4.251e-03 |
| Control | Clinician Consensus *∩* GOLEM | -4.6041 | 4.142e-06 |

Supplementary Table 5. Full Results of the Removing Medications Experiment.

| **Model** | **DAG** | **AUPRC** | **AUROC** | **Precision** | **Recall** | **F1 Score** | **Accuracy** | **ECE** | **# Features** | **# Biomarkers** |
| --- | --- | --- | --- | --- | --- | --- | --- | --- | --- | --- |
| LGBN | Control | 0.76 (0.71, 0.80) | 0.90 (0.88, 0.92) | 0.92 (0.91, 0.94) | 0.78 (0.76, 0.80) | 0.83 (0.81, 0.85) | 0.94 (0.94, 0.95) | 0.137 | 624 | 34 |
| XGB | Control | 0.80 (0.77, 0.83) | 0.92 (0.91, 0.94) | 0.92 (0.90, 0.94) | 0.80 (0.78, 0.82) | 0.85 (0.83, 0.87) | 0.95 (0.94, 0.96) | 0.09 | 624 | 34 |
| LGBN | Correlation | 0.72 (0.68, 0.76) | 0.86 (0.84, 0.89) | 0.94 (0.93, 0.96) | 0.73 (0.71, 0.75) | 0.80 (0.77, 0.82) | 0.94 (0.93, 0.95) | 0.129 | 70 | 26 |
| XGB | Correlation | 0.75 (0.71, 0.79) | 0.89 (0.87, 0.91) | 0.90 (0.88, 0.92) | 0.78 (0.76, 0.80) | 0.82 (0.80, 0.85) | 0.94 (0.93, 0.95) | 0.111 | 70 | 26 |
| LGBN | Clinician Consensus *∪* PC-MB | 0.75 (0.71, 0.78) | 0.90 (0.88, 0.92) | 0.93 (0.91, 0.95) | 0.75 (0.73, 0.78) | 0.81 (0.79, 0.83) | 0.94 (0.93, 0.95) | 0.133 | 198 | 11 |
| XGB | Clinician Consensus *∪* PC-MB | 0.77 (0.74, 0.81) | 0.91 (0.89, 0.93) | 0.91 (0.89, 0.93) | 0.79 (0.77, 0.81) | 0.84 (0.81, 0.86) | 0.94 (0.94, 0.95) | 0.1 | 198 | 11 |
| LGBN | Clinician Consensus *∪* GOLEM | 0.74 (0.70, 0.78) | 0.90 (0.88, 0.92) | 0.93 (0.91, 0.94) | 0.76 (0.73, 0.78) | 0.82 (0.79, 0.84) | 0.94 (0.93, 0.95) | 0.137 | 271 | 15 |
| XGB | Clinician Consensus *∪* GOLEM | 0.77 (0.74, 0.81) | 0.91 (0.89, 0.93) | 0.91 (0.89, 0.92) | 0.80 (0.78, 0.82) | 0.84 (0.82, 0.86) | 0.95 (0.94, 0.95) | 0.099 | 271 | 15 |
| LGBN | GOLEM *∪* PC-MB | 0.74 (0.70, 0.78) | 0.90 (0.88, 0.92) | 0.92 (0.90, 0.94) | 0.75 (0.73, 0.78) | 0.81 (0.79, 0.83) | 0.94 (0.93, 0.95) | 0.138 | 271 | 15 |
| XGB | GOLEM *∪* PC-MB | 0.78 (0.75, 0.82) | 0.91 (0.89, 0.93) | 0.91 (0.89, 0.93) | 0.79 (0.77, 0.82) | 0.84 (0.82, 0.86) | 0.95 (0.94, 0.95) | 0.096 | 271 | 15 |
| LGBN | Clinician Consensus | 0.75 (0.72, 0.79) | 0.90 (0.88, 0.92) | 0.94 (0.93, 0.96) | 0.75 (0.73, 0.77) | 0.81 (0.79, 0.83) | 0.94 (0.93, 0.95) | 0.131 | 160 | 9 |
| XGB | Clinician Consensus | 0.75 (0.72, 0.79) | 0.90 (0.88, 0.92) | 0.91 (0.90, 0.93) | 0.78 (0.76, 0.80) | 0.83 (0.81, 0.85) | 0.94 (0.94, 0.95) | 0.1 | 160 | 9 |
| LGBN | Clinician Consensus *∩* PC-MB | 0.74 (0.70, 0.77) | 0.89 (0.87, 0.91) | 0.94 (0.92, 0.95) | 0.74 (0.72, 0.77) | 0.81 (0.78, 0.83) | 0.94 (0.93, 0.95) | 0.133 | 141 | 8 |
| XGB | Clinician Consensus *∩* PC-MB | 0.76 (0.73, 0.80) | 0.91 (0.89, 0.92) | 0.91 (0.89, 0.93) | 0.78 (0.76, 0.80) | 0.83 (0.81, 0.85) | 0.94 (0.94, 0.95) | 0.1 | 141 | 8 |
| LGBN | PC-MB | 0.74 (0.70, 0.77) | 0.90 (0.88, 0.92) | 0.93 (0.91, 0.95) | 0.75 (0.72, 0.77) | 0.81 (0.78, 0.83) | 0.94 (0.93, 0.95) | 0.134 | 179 | 10 |
| XGB | PC-MB | 0.78 (0.75, 0.81) | 0.91 (0.89, 0.93) | 0.92 (0.90, 0.93) | 0.80 (0.77, 0.82) | 0.84 (0.82, 0.86) | 0.95 (0.94, 0.95) | 0.101 | 179 | 10 |
| LGBN | GOLEM | 0.72 (0.68, 0.76) | 0.89 (0.87, 0.91) | 0.92 (0.90, 0.94) | 0.74 (0.72, 0.76) | 0.80 (0.78, 0.82) | 0.94 (0.93, 0.95) | 0.136 | 180 | 10 |
| XGB | GOLEM | 0.78 (0.75, 0.81) | 0.92 (0.90, 0.93) | 0.91 (0.89, 0.93) | 0.79 (0.77, 0.82) | 0.84 (0.82, 0.86) | 0.95 (0.94, 0.95) | 0.098 | 180 | 10 |
| LGBN | GOLEM *∩* PC-MB | 0.71 (0.67, 0.76) | 0.89 (0.87, 0.91) | 0.93 (0.91, 0.94) | 0.74 (0.71, 0.76) | 0.80 (0.77, 0.82) | 0.94 (0.93, 0.95) | 0.131 | 88 | 5 |
| XGB | GOLEM *∩* PC-MB | 0.76 (0.73, 0.80) | 0.90 (0.89, 0.92) | 0.90 (0.88, 0.92) | 0.79 (0.77, 0.81) | 0.83 (0.81, 0.85) | 0.94 (0.94, 0.95) | 0.102 | 88 | 5 |
| LGBN | Clinician Consensus *∩* GOLEM | 0.72 (0.67, 0.75) | 0.88 (0.86, 0.90) | 0.94 (0.92, 0.95) | 0.73 (0.70, 0.75) | 0.79 (0.77, 0.81) | 0.94 (0.93, 0.94) | 0.13 | 69 | 4 |
| XGB | Clinician Consensus *∩* GOLEM | 0.74 (0.70, 0.77) | 0.89 (0.87, 0.91) | 0.89 (0.87, 0.91) | 0.78 (0.76, 0.80) | 0.82 (0.80, 0.84) | 0.94 (0.93, 0.95) | 0.106 | 69 | 4 |

Supplementary Table 6. Results *of Delong’s test 72 for XGBoost models in the Only Vitals and Labs experiments.*

**Model 1	Model 2	Z-score	P-value**

| Control | Correlation | -3.4961 | 4.721e-04 |
| --- | --- | --- | --- |
| Control | Clinician Consensus *∪* PC-MB | -1.8281 | 6.754e-02 |
| Control | Clinician Consensus *∪* GOLEM | -0.7281 | 4.665e-01 |
| Control | GOLEM *∪* PC-MB | -1.1787 | 2.385e-01 |
| Control | Clinician Consensus | -4.025 | 5.698e-05 |
| Control | Clinician Consensus *∩* PC-MB | -4.025 | 5.698e-05 |
| Control | PC-MB | -1.8281 | 6.754e-02 |
| Control | GOLEM | -1.7074 | 8.774e-02 |
| Control | GOLEM *∩* PC-MB | -3.8557 | 1.154e-04 |

Supplementary Table 7. Full Results of the Only Vitals and Labs experiments.

| **Model** | **DAG** | **AUPRC** | **AUROC** | **Precision** | **Recall** | **F1 Score** | **Accuracy** | **ECE** | **# Features** | **# Biomarkers** |
| --- | --- | --- | --- | --- | --- | --- | --- | --- | --- | --- |
| LGBN | Control | 0.75 (0.71, 0.79) | 0.90 (0.87, 0.92) | 0.92 (0.90, 0.94) | 0.77 (0.75, 0.80) | 0.83 (0.81, 0.85) | 0.94 (0.94, 0.95) | 0.137 | 521 | 28 |
| XGB | Control | 0.78 (0.75, 0.82) | 0.92 (0.90, 0.93) | 0.90 (0.88, 0.92) | 0.80 (0.78, 0.82) | 0.84 (0.82, 0.86) | 0.95 (0.94, 0.95) | 0.093 | 521 | 28 |
| LGBN | Correlation | 0.69 (0.66, 0.74) | 0.85 (0.82, 0.87) | 0.94 (0.93, 0.96) | 0.72 (0.70, 0.74) | 0.78 (0.76, 0.81) | 0.94 (0.93, 0.94) | 0.13 | 53 | 20 |
| XGB | Correlation | 0.75 (0.71, 0.78) | 0.89 (0.87, 0.91) | 0.90 (0.88, 0.92) | 0.78 (0.76, 0.80) | 0.83 (0.80, 0.85) | 0.94 (0.93, 0.95) | 0.114 | 53 | 20 |
| LGBN | Clinician Consensus *∪* PC-MB | 0.69 (0.65, 0.74) | 0.87 (0.85, 0.90) | 0.91 (0.89, 0.93) | 0.73 (0.70, 0.75) | 0.78 (0.76, 0.81) | 0.93 (0.93, 0.94) | 0.145 | 131 | 7 |
| XGB | Clinician Consensus *∪* PC-MB | 0.76 (0.73, 0.80) | 0.91 (0.89, 0.92) | 0.89 (0.87, 0.91) | 0.79 (0.77, 0.81) | 0.83 (0.81, 0.85) | 0.94 (0.93, 0.95) | 0.104 | 131 | 7 |
| LGBN | Clinician Consensus *∪* GOLEM | 0.72 (0.68, 0.75) | 0.88 (0.86, 0.90) | 0.90 (0.88, 0.93) | 0.73 (0.71, 0.75) | 0.79 (0.76, 0.81) | 0.93 (0.93, 0.94) | 0.142 | 204 | 11 |
| XGB | Clinician Consensus *∪* GOLEM | 0.77 (0.74, 0.81) | 0.91 (0.90, 0.93) | 0.91 (0.89, 0.93) | 0.79 (0.77, 0.81) | 0.84 (0.82, 0.86) | 0.95 (0.94, 0.95) | 0.101 | 204 | 11 |
| LGBN | GOLEM *∪* PC-MB | 0.72 (0.68, 0.76) | 0.88 (0.86, 0.90) | 0.91 (0.89, 0.93) | 0.73 (0.71, 0.76) | 0.79 (0.77, 0.81) | 0.93 (0.93, 0.94) | 0.142 | 223 | 12 |
| XGB | GOLEM *∪* PC-MB | 0.77 (0.73, 0.80) | 0.91 (0.89, 0.93) | 0.91 (0.89, 0.93) | 0.78 (0.76, 0.81) | 0.83 (0.81, 0.85) | 0.94 (0.94, 0.95) | 0.098 | 223 | 12 |
| LGBN | Clinician Consensus | 0.68 (0.64, 0.72) | 0.87 (0.84, 0.89) | 0.92 (0.90, 0.94) | 0.70 (0.68, 0.72) | 0.76 (0.73, 0.78) | 0.93 (0.92, 0.94) | 0.146 | 93 | 5 |
| XGB | Clinician Consensus | 0.73 (0.69, 0.76) | 0.89 (0.87, 0.91) | 0.89 (0.87, 0.91) | 0.78 (0.75, 0.80) | 0.82 (0.80, 0.84) | 0.94 (0.93, 0.95) | 0.108 | 93 | 5 |
| LGBN | Clinician Consensus *∩* PC-MB | 0.68 (0.64, 0.72) | 0.87 (0.84, 0.89) | 0.92 (0.90, 0.94) | 0.70 (0.68, 0.72) | 0.76 (0.73, 0.78) | 0.93 (0.92, 0.94) | 0.146 | 93 | 5 |
| XGB | Clinician Consensus *∩* PC-MB | 0.73 (0.69, 0.76) | 0.89 (0.87, 0.91) | 0.89 (0.87, 0.91) | 0.78 (0.75, 0.80) | 0.82 (0.80, 0.84) | 0.94 (0.93, 0.95) | 0.108 | 93 | 5 |
| LGBN | PC-MB | 0.69 (0.65, 0.74) | 0.87 (0.85, 0.90) | 0.91 (0.89, 0.93) | 0.73 (0.70, 0.75) | 0.78 (0.76, 0.81) | 0.93 (0.93, 0.94) | 0.145 | 131 | 7 |
| XGB | PC-MB | 0.76 (0.73, 0.80) | 0.91 (0.89, 0.92) | 0.89 (0.87, 0.91) | 0.79 (0.77, 0.81) | 0.83 (0.81, 0.85) | 0.94 (0.93, 0.95) | 0.104 | 131 | 7 |
| LGBN | GOLEM | 0.69 (0.65, 0.73) | 0.87 (0.85, 0.89) | 0.91 (0.89, 0.93) | 0.72 (0.70, 0.74) | 0.78 (0.75, 0.80) | 0.93 (0.92, 0.94) | 0.142 | 149 | 8 |
| XGB | GOLEM | 0.76 (0.72, 0.79) | 0.91 (0.89, 0.92) | 0.90 (0.88, 0.92) | 0.79 (0.77, 0.81) | 0.83 (0.81, 0.85) | 0.94 (0.94, 0.95) | 0.102 | 149 | 8 |
| LGBN | GOLEM *∩* PC-MB | 0.60 (0.56, 0.65) | 0.85 (0.83, 0.87) | 0.89 (0.86, 0.91) | 0.67 (0.65, 0.69) | 0.72 (0.70, 0.75) | 0.92 (0.91, 0.93) | 0.15 | 57 | 3 |
| XGB | GOLEM *∩* PC-MB | 0.71 (0.68, 0.75) | 0.89 (0.87, 0.91) | 0.86 (0.84, 0.89) | 0.77 (0.75, 0.80) | 0.81 (0.79, 0.83) | 0.93 (0.93, 0.94) | 0.113 | 57 | 3 |
| LGBN | Clinician Consensus *∩* GOLEM | 0.56 (0.51, 0.60) | 0.82 (0.79, 0.84) | 0.87 (0.84, 0.90) | 0.63 (0.60, 0.65) | 0.67 (0.64, 0.70) | 0.91 (0.90, 0.92) | 0.152 | 38 | 2 |
| XGB | Clinician Consensus *∩* GOLEM | 0.66 (0.62, 0.70) | 0.86 (0.84, 0.88) | 0.85 (0.82, 0.87) | 0.74 (0.72, 0.76) | 0.78 (0.76, 0.80) | 0.93 (0.92, 0.93) | 0.116 | 38 | 2 |

**Supplemental Figures**

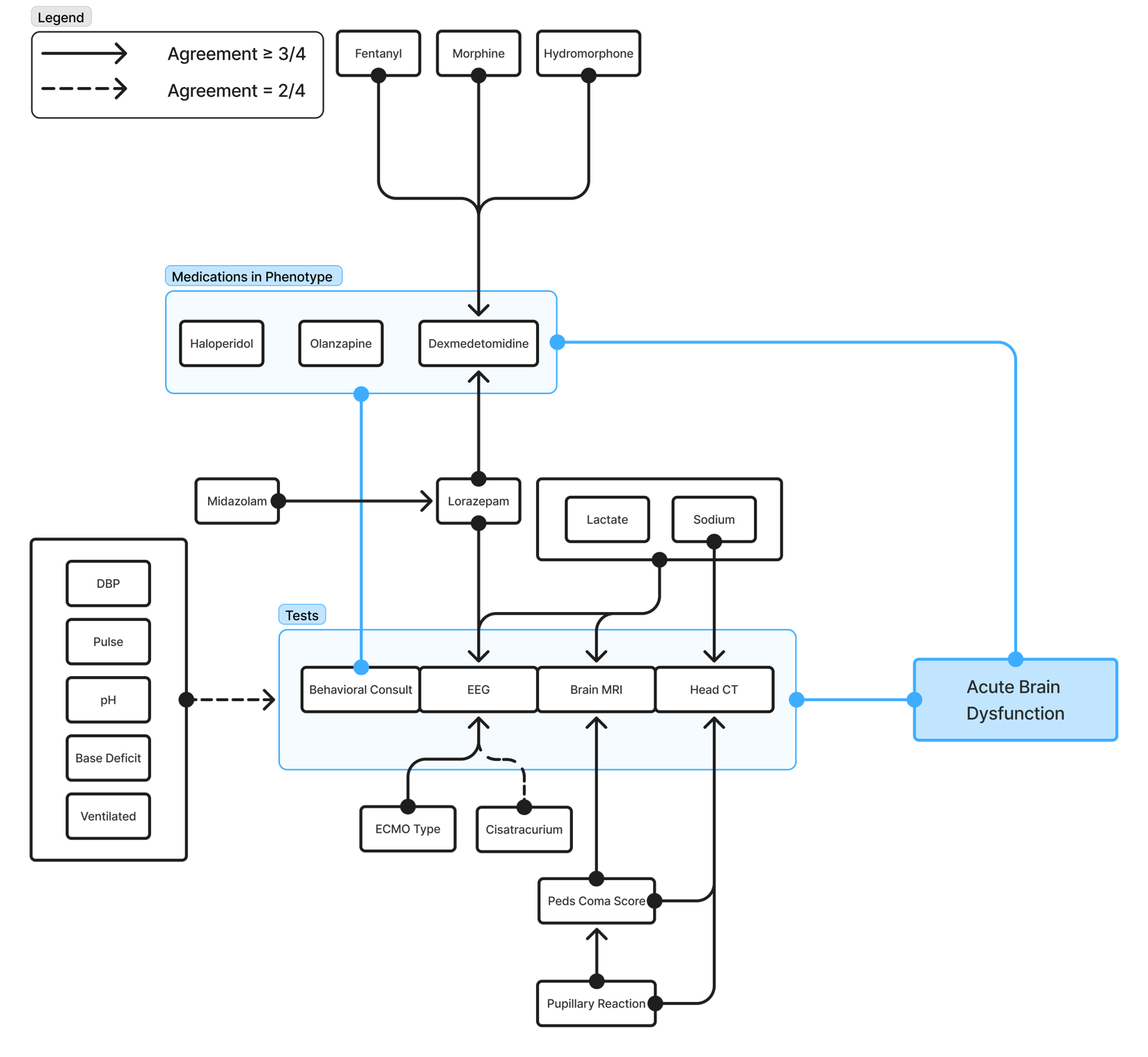

Supplemental Figure 1. Clinician’s Consensus DAG. Blue lines indicate components of the Acute Brain Dysfunction (ABD) computable phenotype. Solid black lines represent edges added by at least 3*/*4 of clinicians. Dashed lines indicate edges added by at least 2*/*4 of clinicians. Some biomarkers were joined in a group to minimize overlapping edges. The blue lines from Behavioral Consult to “Medications in the Phenotype” indicate that the presence of those medications will only count towards the phenotype if they’re given after a Behavioral Consult.

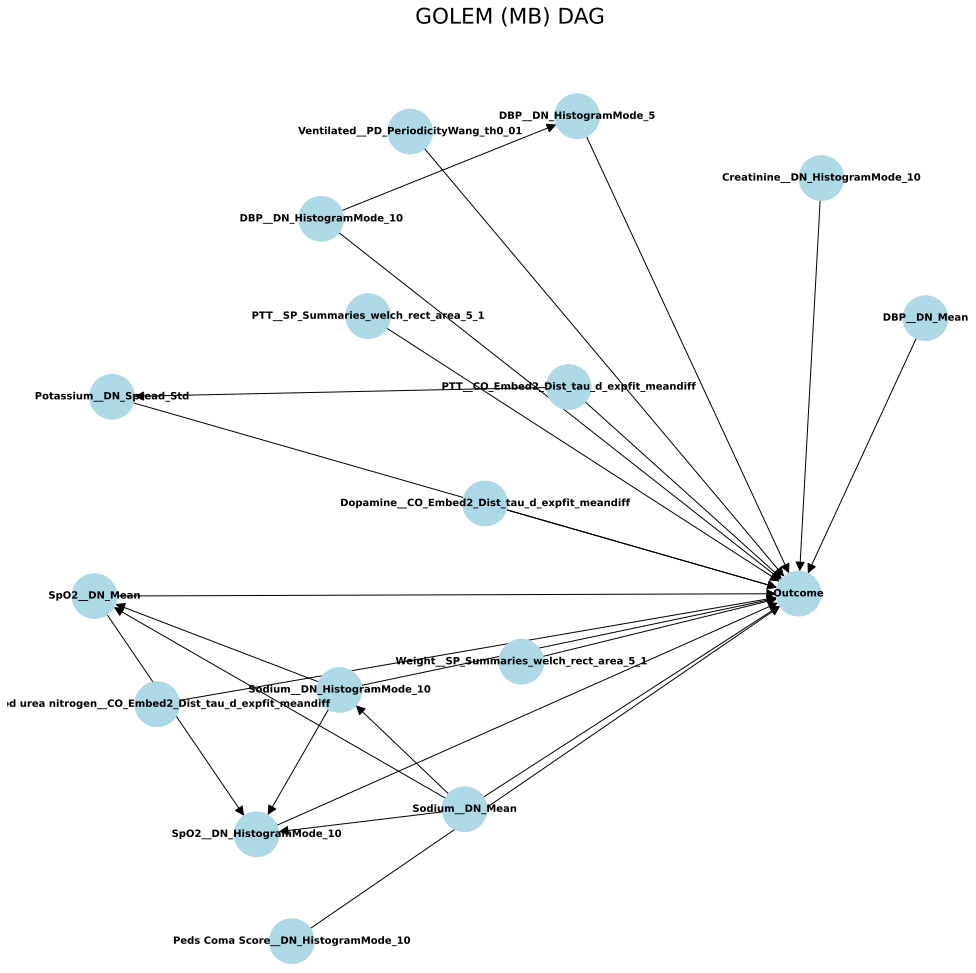

Supplemental Figure 2. Markov Blanket of the GOLEM DAG. Edges represent the direction of potential causes. In this version of the DAG, we show interactions across parents of our target Outcome (Acquired ABD). We also show which feature and biomarker combinations were identified as potential causes. See the [official documenta-tion](https://time-series-features.gitbook.io/catch22/information-about-catch22/feature-descriptions) ^71^ for an in-depth description of Catch22 features.

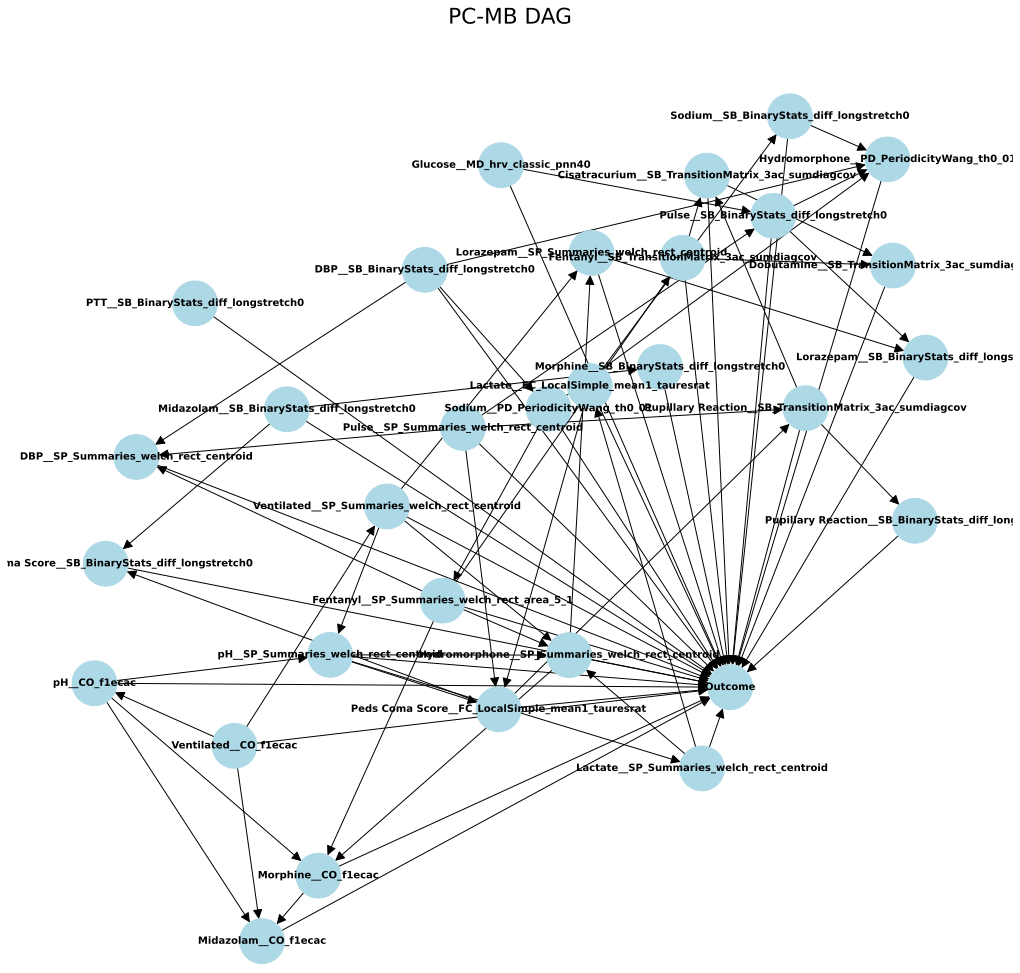

Supplemental Figure 3. PC-MB DAG. Edges represent the direction of potential causes. In this version of the DAG, we show interactions across parents of our target Outcome (Acquired ABD). We also show which feature and biomarker combinations were identified as potential causes. See the [official documentation](https://time-series-features.gitbook.io/catch22/information-about-catch22/feature-descriptions) ^71^ for an in-depth description of Catch22 features.

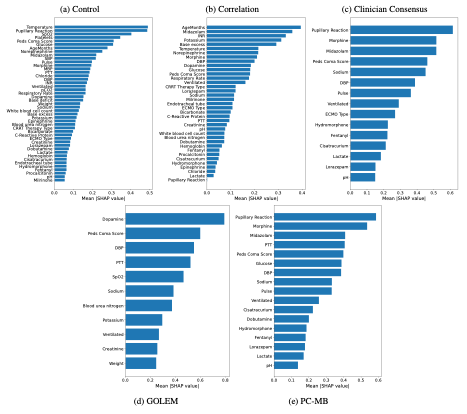

Supplemental Figure 4. Biomarker importance across XGBoost models for the Biomarker Selection Experiments.

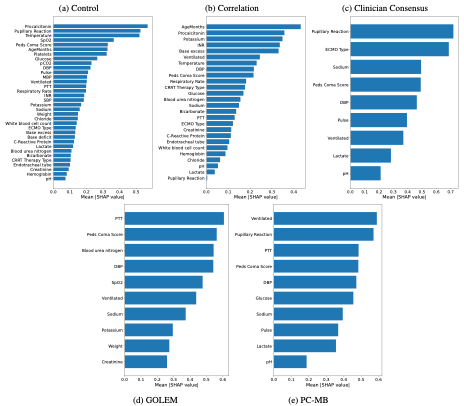

Supplemental Figure 5. Biomarker importance across XGBoost models for the No Medications Experiments.

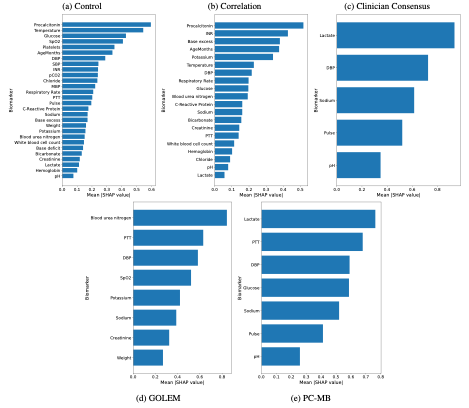

*Supplemental Figure 6. Biomarker importance across XGBoost models for the Only Vitals and Labs experiments.*

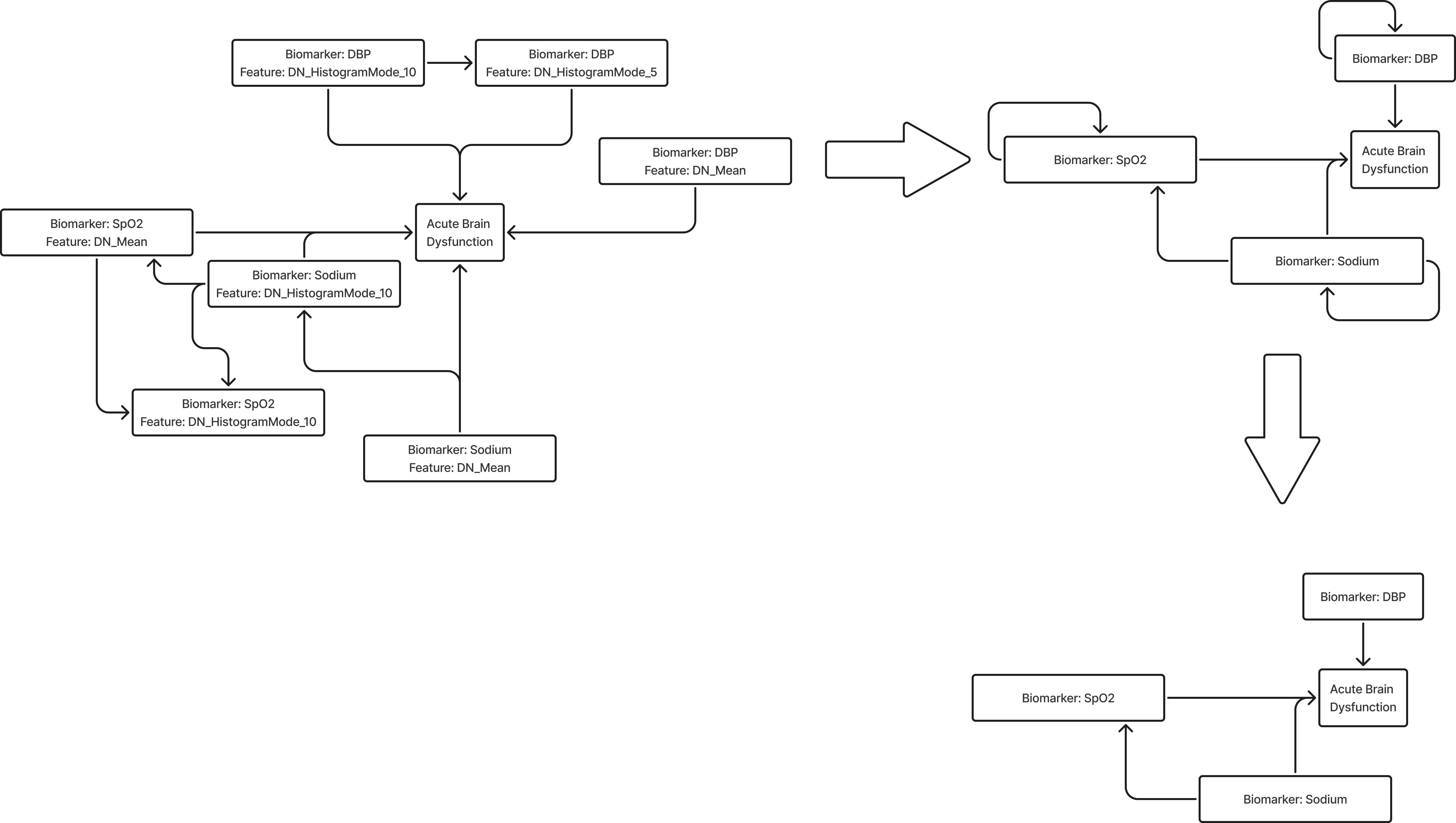

Supplemental Figure 7. Illustration of how we simplified the PC-MB and GOLEM DAGs. Features belonging to the same biomarker were collapsed while maintaining all edges. Then self-loops were removed.
